# An emerging role of microplastics in the etiology of lung ground glass nodules

**DOI:** 10.1101/2021.04.22.21255586

**Authors:** Qiqing Chen, Jiani Gao, Hairui Yu, Hang Su, Yan Yang, Yajuan Cao, Qun Zhang, Yijiu Ren, Huahong Shi, Chang Chen, Haipeng Liu

**Affiliations:** State Key Laboratory of Estuarine and Coastal Research, East China Normal University, Shanghai 200241, China; Department of Thoracic Surgery, Shanghai Pulmonary Hospital, Tongji University, Shanghai 200433, China; Clinical and Translational Research Center, Shanghai Pulmonary Hospital, Tongji University, Shanghai 200433, China; Central Laboratory, Shanghai Pulmonary Hospital, Tongji University, Shanghai 200433, China

## Abstract

Pulmonary ground glass nodules (GGNs) have been increasingly identified in past decades and is becoming an important clinical dilemma in oncology. Meanwhile, humans persistently inhale microplastics which are dominant in the air. However, the retention of “non-self” microplastics in human lung and its correlation with pulmonary GGNs remains elusive. In this study, we firstly demonstrated the presence of microfibers and microplastics in human lung, with higher detection rates in GGNs in comparison to those in normal tissue. Moreover, both types and colors of microfibers in tumor were richer than those in normal tissues. Intriguingly, high risk of microfibers exposure predisposes the formation of pulmonary GGN. Further, increased roughness surface was observed in microfibers isolated in human lung, indicating the possible link of surface roughness to the formation of pulmonary GGN. Collectively, our findings reveal an emerging role of environmental microplastics exposure in the etiology of pulmonary GGN.

**One Sentence Summary:** The exposure of environmental microplastics is a risk factor of pulmonary GGN.

## Introduction

Microplastic was firstly proposed as a marine environmental problem, but with its widespread occurrence in freshwater and estuary^1-3^, in land and mountains^4, 5^, and in glaciers and polar regions^6, 7^, microplastic pollution has become a global health issue and aroused some debates among scientists^8-10^. Recently, the pervasiveness of microplastics has also been verified in both indoor and outdoor air environments^11^, and the abundance of microplastics in the air is one order of magnitude higher than that in other media^12^. This means all living animals breathing with lungs (including humans) cannot escape the fate of inhaling microplastics.

Though the inhalation of mineral fiber such as asbestos has been widely recognized^13, 14^, whereas pulmonary retention of nonmineral fibers including artificial fibrous microplastics or natural cotton fibers which are difficult to be degraded due to their stable structure remains unclear. The amount of microplastics humans take in through respiration may be much higher than other routes since microplastics are dominant in the air^12^. It has been predicted that the amount of microplastics inhaled by human body through breathing was dozens to hundreds per day^15, 16^. An early preliminary study reported the presence of fibers in human lung; however, there was a lack of detailed characterization of the fibers including the size and type, color and morphology^17^. Of note, there is no direct evidence to demonstrate what type and abundance of microplastics exist in lung tissue. In addition, whether the retention of microplastics and the long-term friction between microplastics and lung tissue are related to some respiratory diseases including lung cancer is largely unknown.

Pulmonary ground glass nodules (GGNs) are areas of lesions of homogenous density and with hazy increase in density in the lung field that does not obscure the broncho vascular structure as identified on low-dose computed tomography (LDCT)^18-20^. Their etiology is broad and the presumed significance is highly dependent on the underlying disease context^21, 22^. In addition to diagnostic techniques to increase the detection rate, the main causes of GGNs include genetic factors and gender factors^23-26^, but research suggests that environmental factors may affect the occurrence of GGNs or lung cancer^24, 27, 28^. When exposed to asbestos, vinyl chloride, or other environmental factors, these substances can be inhaled into the lungs to mount an immune response. The inflammatory response may form GGNs by wrapping, organizing or forming granulomas^27^. In addition, a long time exposure to a dusty environment without proper protection has been reported to impair lung function and cause GGNs^28^. Moreover, people who maintain smoking habits for a long time are more likely to cause GGNs^24,29^. The proportion of plastics produced and used in the current industrial civilization has increased year by year, leading to the existence of a large number of microplastics in the environment. This trend is consistent with the increasing incidence of GGN in recent years. In this study, we aimed to systematically detect and characterize the microplastics in GGNs and adjacent lung tissues, and analyzed the correlation of the presence of microplastics with the occurrence of GGNs.

## Results

### Identification of microfibers in human lung tissue of pulmonary GGNs patients

To detect whether microfibers are present in human lung tissues, we collected surgically dissected human lung tissues including GGNs and adjacent normal tissues in patients with pulmonary GGNs which are pathologically diagnosed. Pulmonary GGN can be observed in preinvasive lesions such as atypical adenomatous hyperplasia (AAH), adenocarcinoma in situ (AIS), or in malignancies such as minimally invasive adenocarcinoma (MIA), lepidic-predominant invasive adenocarcinomas (LPA)^30^. By using hydrogen peroxide digestion method, we successfully isolated microfibers from the lung tissues in both tumor (**Figure 1a-e**) and normal (**Figure 1f-h**) tissues. The most common types of microfibers are cotton **(Figure 1a)**, rayon **(Figure 1b, g)**, polyester **(Figure 1c, h)**, denim **(Figure 1f)**, and the representative examples are presented. Besides, we also found some kind of microfibers that rarely found in water or soil, such as phenoxy resin (**Figure 1d**) and chipboard (**Figure 1e**). There was no obvious difference in gross morphology between the tumor and normal groups.

**Fig. 1.**
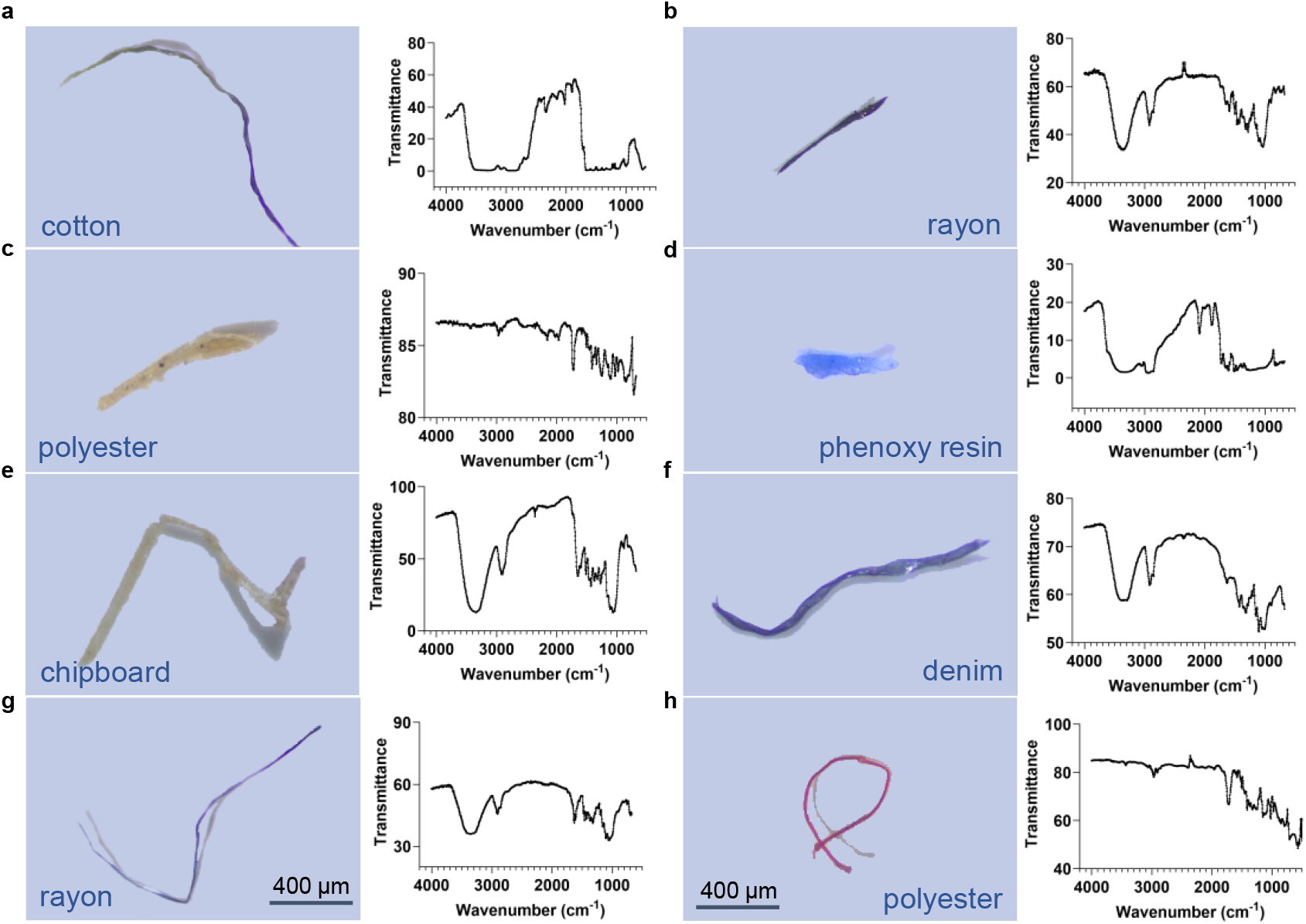
Identified of microfibers (including microplastics) in human lung tissues. Images under microscope and infrared spectrum of (a) cotton in tumor tissue; (b) rayon in tumor tissue; (c) polyester in tumor tissue; (d) phenoxy resin in tumor tissue; (e) chipboard in tumor tissue; (f) denim in normal tissue; (g) rayon in normal tissue; (h) polyester in normal tissue.

### *In situ* detection of microfibers in human lung tissue of pulmonary GGNs patients

To further demonstrate the presence of microfibers in human lung, we employed the Laser Direct Infrared (LDIR) method to detect the microfibers in the lung tissue slice from pulmonary GGNs patients *in situ*. By analyzing cryo-sectioned slices, we successfully observed the presence of a microfiber (width 24 μm, length 887 μm) in the lung tissue slice by microscopy observation with both visible light (**Figure 2a, d**) and infrared spectrum (**Figure 2b, c**). The infrared absorption spectrum revealed that the composition of the microfiber was cellulose **(Figure 2e)**. This was further consolidated by the collected signals with the specific cellulose characteristics **(Figure 2f)**, which indicates that this fiber component is indeed cellulose and is different from the surrounding components. As the characteristic signal intensities of this microfiber were uneven at different regions **(Figure 2f)**, we suppose that this microfiber should be embedded in the tissue. We therefore for the first time provide solid *in situ* evidence to support the presence of microfiber in human lung tissue.

**Fig. 2.**
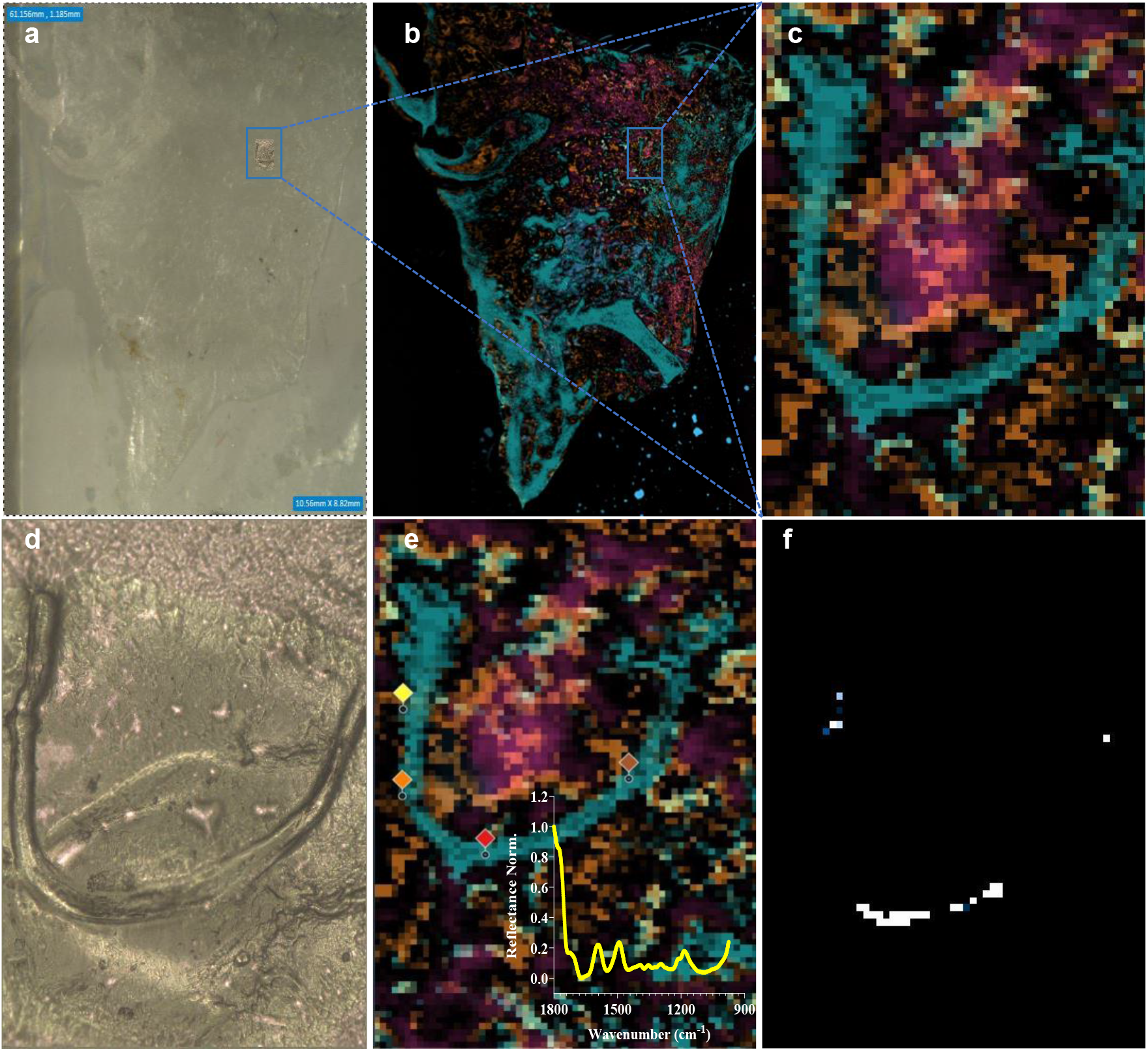
*In situ* detection of a cellulose microfiber in the tumor lung tissue slice. (a) the whole tissue section image under vis-spectrum; (b) the whole tissue section image under IR-spectrum; (c) microfiber detected under IR-spectrum; (d) microfiber detected under vis-spectrum; (e) detected dots with confirmed cellulose composition; (f) strong signals (in white color) with the specific cellulose characteristics.

### Higher detection rate of microfibers in the GGNs

To understand whether the presence of microfibers is associated with the occurrence of GGNs, we further compared the frequency of the detected microfibers in the GGNs and adjacent normal lung tissues. In 50 pairs of samples, microfibers have been found in 29 tumor and 23 normal tissues (**Figure 3a**), in the female subgroup, the detection rates of microplastics in tumor and normal tissues were different (**Figure 3c-3d**), thus, the positive detection rates were 58% and 46% for tumor and normal tissues, respectively. Moreover, a total of 38 microfibers were detected in tumor tissues, accounting for 58.46% of the total detected microfibers; while 27 microfibers were detected in normal tissues, accounting for 41.54% (**Figure 3b**). Among the detected microfibers, 24 were microplastics, constituting 36.92% of the total microfibers **(Figure 3b)**. Sixteen microplastics were detected in the tumor tissues, which was twice than that of the normal tissues **(Figure 3b)**. Taken together, in comparison to that in normal lung tissues, the detection rate of microfibers or microplastics in GGNs was significantly higher, indicating a possible link of the presence of microfibers to the occurrence of GGNs.

**Fig. 3.**
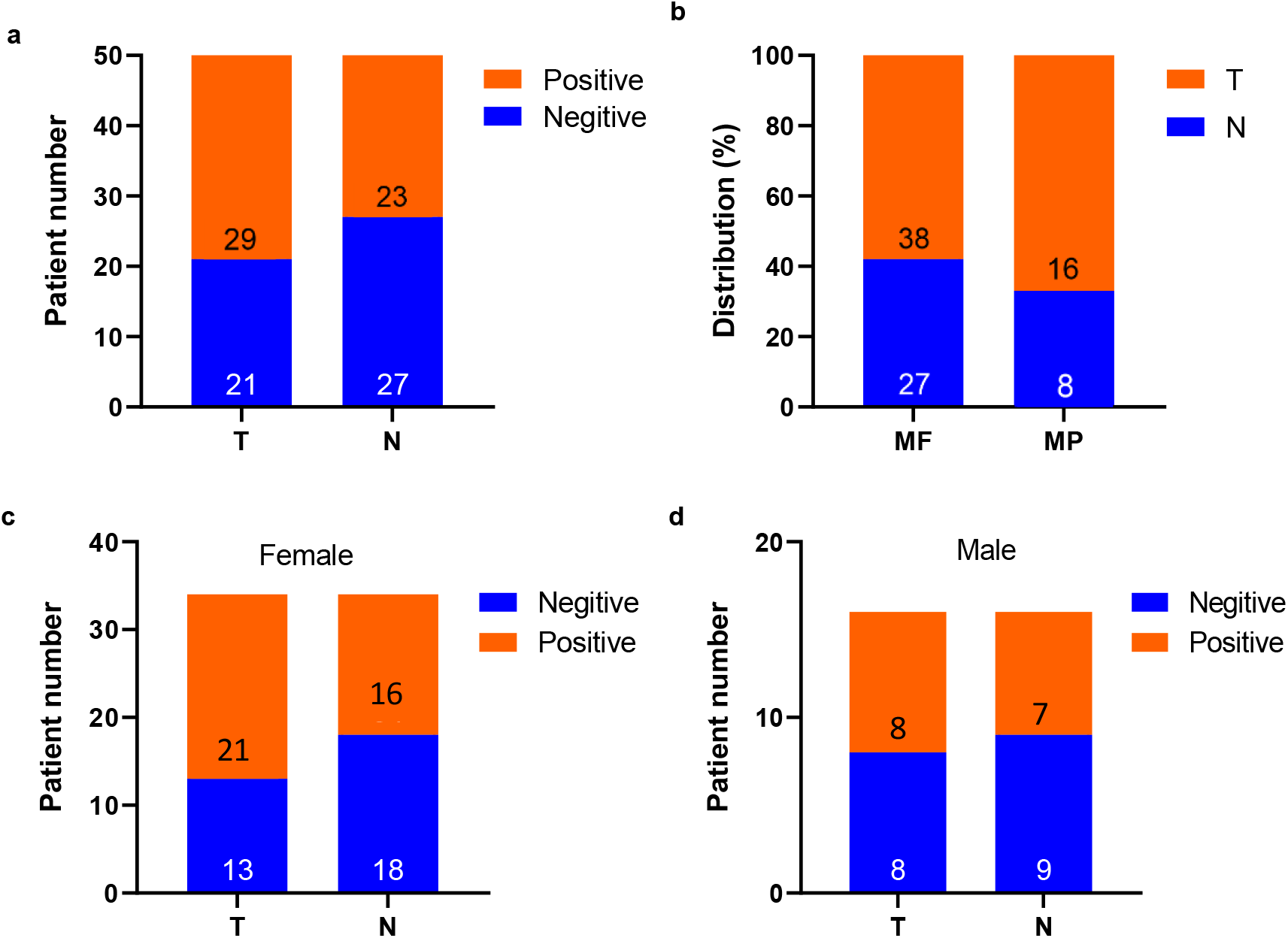
Microfiber and microplastic tissue distribution and detection frequency. (a) Patient numbers with or without microfibers detected in lungs; (b) Distribution of microfibers and microplastics. T: tumor tissue; N: normal tissue; MF: microfiber; MP: microplastic; (c) Patient numbers with or without microfibers detected in women; (d) Patient numbers with or without microfibers detected in men. The annotated values on each bar of figures a, c, and d represent patient numbers; the annotated values on each bar of figure b represent detected microfiber or microplastic numbers.

### Characterization of microfibers in the lung tissues of pulmonary GGN patients

Next, we checked the characteristics including width, length, type and color of microfibers isolated in human lung tissues. The average values of length (1.45±0.98 mm) and width (35.74±21.09 μm) of microfibers in tumor tissues are slightly higher than those in the normal (1.38±0.96 mm for length, and 32.81±16.91 μm for width), but there is no significant difference between them **(Figure 4a)**. Additionally, both the length of microplastics in tumor (1.75±0.79 mm) and normal (1.49±0.96 mm) tissues, and the width of microplastics in tumor (34.29±18.60 μm) and normal (34.15±17.91 μm) tissues are quite similar **(Figure 4a)**. Moreover, microfibers detected in the lungs were mainly >1000 μm in length, with 63% for microfiber and 50% for microplastic in tumor tissue, 48% for microfiber and 63% for microplastic in normal tissue (**Figure S1a-b**). The width of microfiber mainly falls in <30 μm in both tumor and normal tissues, accounting 47.37% and 51.85%, respectively. However, microplastic widths have the highest proportion of 30-50 μm in tumor tissue, accounting 56.25%, while the highest proportions of < 30 and 30-50 in normal tissue are both 37.5% (**Figure S1c-d**).

**Fig. 4.**
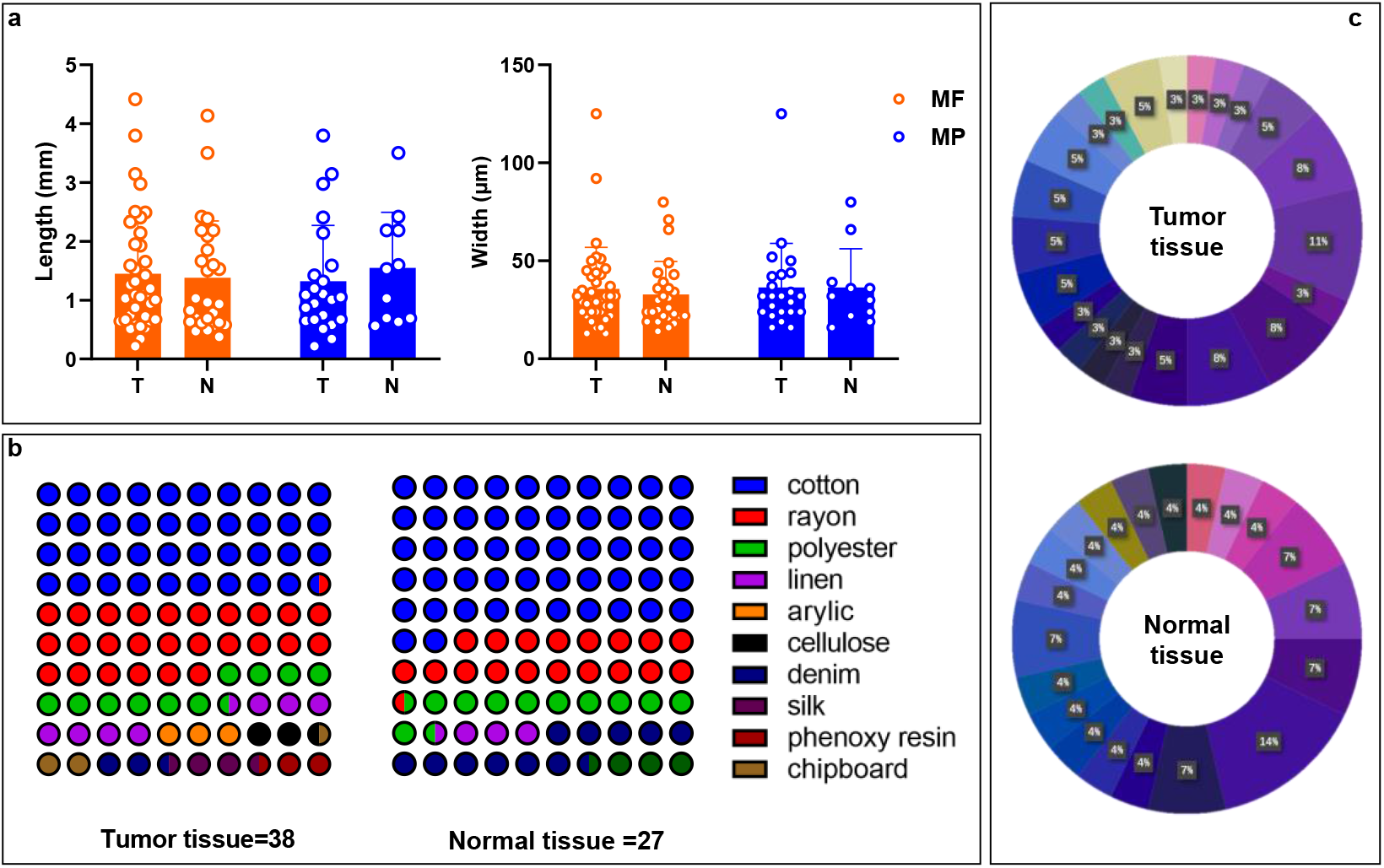
Microfiber characteristics in lung tumor and normal tissues. (a) width and length distribution; (b) composition type distribution (each dot represents 1% of the total microfiber composition); (c) color distribution. T: tumor tissue; N: normal tissue. MF: microfiber; MP: microplastic.

Moreover, multiple types of microfibers were detected in the lung. The dominant type of microfibers is cotton which accounts for 39.47% in tumor and 51.85% in normal tissues, respectively. Rayon ranks the second and constitutes 26.32% and 18.52% of tumor and normal tissues, respectively. The third is polyester, which accounts for 10.53% of tumor and 11.11% of normal tissues. Moreover, 10 kinds of microfibers are detected in tumor, which is more abundant than normal tissues (6 kinds) **(Figure 4b)**. The colors of microfibers, whether in tumor or normal tissues, are mainly purple and blue with different shades, and a small amount of transparent and yellow microfibers are detected. There are more reddish microfibers in normal tissues **(Figure 4c)**.

### Characterization of microfibers in matched tumor and normal lung tissues

Parameters commonly used to describe the characteristics of microfibers including polymer composition, size, color, density, shape^31^. To further make clear which characteristic of microfibers may be responsible for the occurrence of GGNs, we also conducted matched samples comparison for the case that microfibers were detected in both tumor and normal tissues of the same patient. Although the mean values of width and length in tumor are higher than those in normal tissues, the difference is not significant **(Figure 5a)**, and only one tenth of the colors and one third of the fiber composition types are the same in matched tumor and normal tissues **(Figure 5b-c)**.

**Fig. 5.**
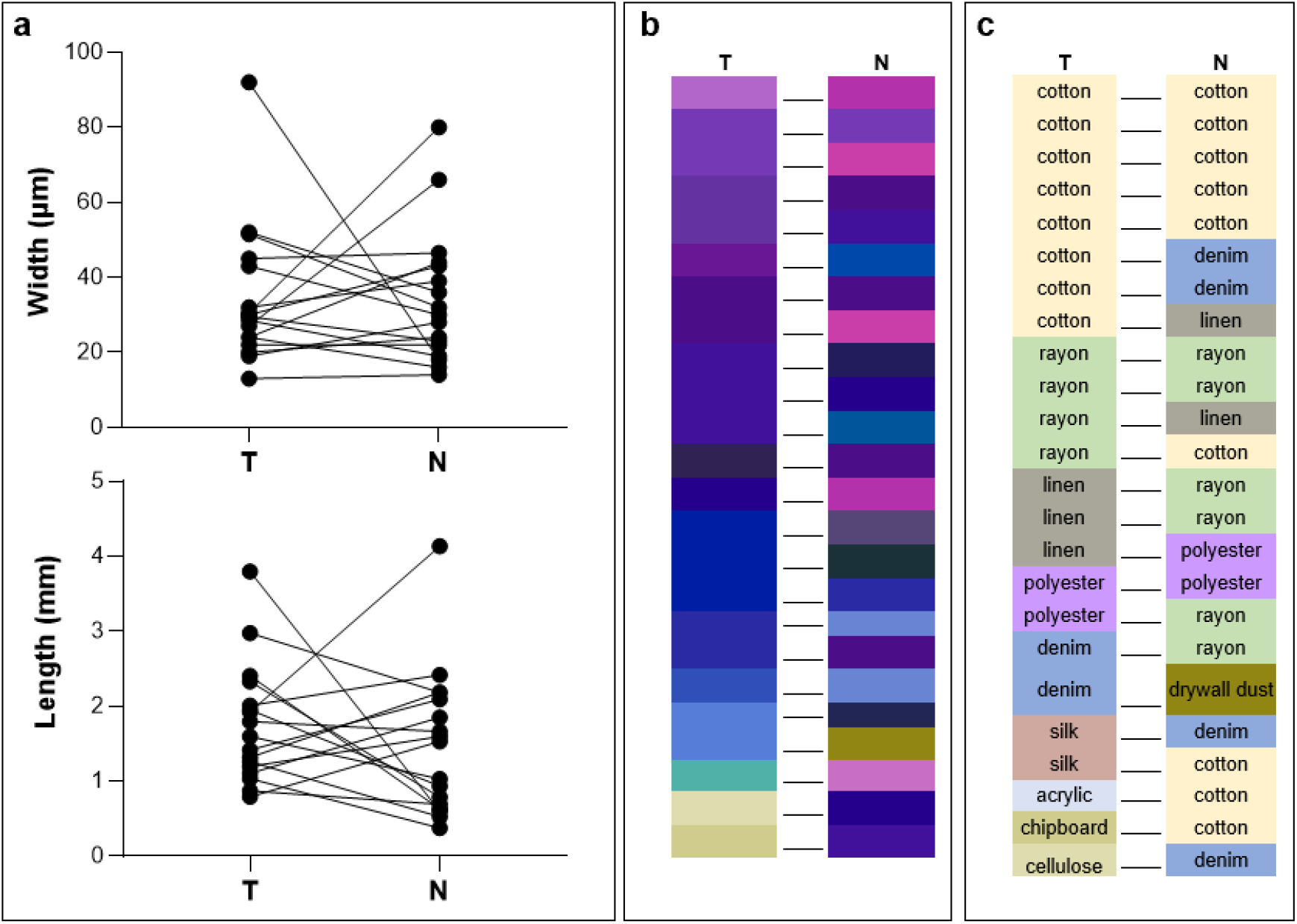
Microfiber characteristics in matched tumor/normal samples. (a) width and length distribution; (b) color and composition type distribution. T: tumor tissue; N: normal tissue. Note: Only patients samples possessing microfibers in both tumor and adjacent normal tissue are included in this figure.

### High risk of microfibers exposure correlates with the occurrence of pulmonary GGNs

To further study the correlation of microfibers with the occurrence of GGNs, we examined the factors contributing to the accumulation of microfibers in the GGN tumor tissues. Chi-square test revealed that microfibers are more likely to be detected in the tumor tissue if one has a history of high microfiber exposure risk in life or work, with a microfiber detection rate reaches 72% **(Figure 6a)**. However, this phenomenon was not found in the normal tissue, and the detection rate was 45% **(Figure 6a)**. This suggests that the exposure to microfibers may lead to the accumulation of microfibers in the lung, which finally contributes to the formation of GGNs.

**Fig. 6.**
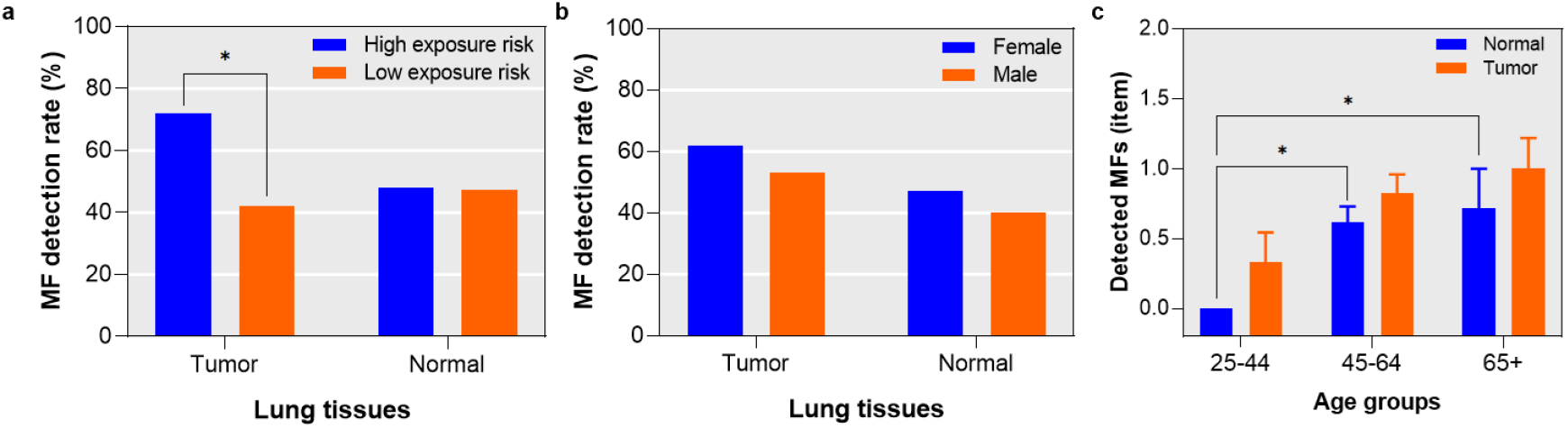
Correlation of microfibers with clinical indices including (a) microfiber exposure history; (b)gender; and (c) age.

Initially, a sex-specific difference in incidence of lung cancer has been reported, women are most likely developing GGN^32, 33^. We next examined whether gender is related to the detection rate of microfibers in the tumor or normal tissue of the lung. The microfiber detection rate of the female (n=34) in either tumor (61.76%) or normal (47.06%) tissues is higher than that of the male (n=16, 53.33% and 40.00%, respectively), but there is no significant difference between the two genders according to the chi-square test result **(Figure 6b)**.

In addition, according to the age classification of lung cancer epidemiology, we found that with the increase of age, the content of microfiber in lung tissue gradually increased **(Figure 6c)**. Importantly, the amount of microfibers in normal tissue is significantly increased with age; however, the magnitude of microfibers in tumor tissues of patients with GGN of all ages are high and not significantly different **(Figure 6c)**. Of note, microfibers were identified in the tumor tissue but not in the any normal tissue of pulmonary GGO patients aged from 25-44 (n=6), indicating that inhalation and accumulation of microfibers in the young people may be a risk factor for the occurrence of GGN.

### Microfibers isolated from lung tissues exhibit high surface roughness

We further characterize the microfibers in human lung tissues by scan electron microscope, the data demonstrate that there are numerous wear and gullies on microfibers as detected by their typical main body structure **(Figure 7a, c, e)** and local surface morphology **(Fig. 7b, d, f)**. Of note, little attention has been paid to the index of surface roughness. This kind of morphology is different from that found in the atmospheric fallout in Shanghai, as we find that there are both worn or unworn microfibers in the air (**Figure S2**), which may be related to their aging degree in the environment. Besides, our data confirmed that the sampling, digestion and identification processes would not be the cause of surface wear **(Figure S3)**. Of note, microfibers in both tumor and normal tissues exhibited a higher roughness value (Ra=0.90±0.84 μm, and 0.79±0.43 μm, respectively) than those in air (Ra=0.38±0.22 μm) (*p*=0.183) (**Figure 7g**), and obvious rougher surface of microfibers in lung tissues than that in the air can be observed (**Figure 7h-k**). Moreover, energy-dispersive X-ray spectroscopy (EDS) revealed that microfibers isolated in the lung did not harbor obvious heavy metal residues (**Figure S4**). Therefore, it is very possible that the surface roughness of microfibers was formed after entering the lungs, formed during microfibers’ long-term interaction with the lungs.

**Figure 7.**
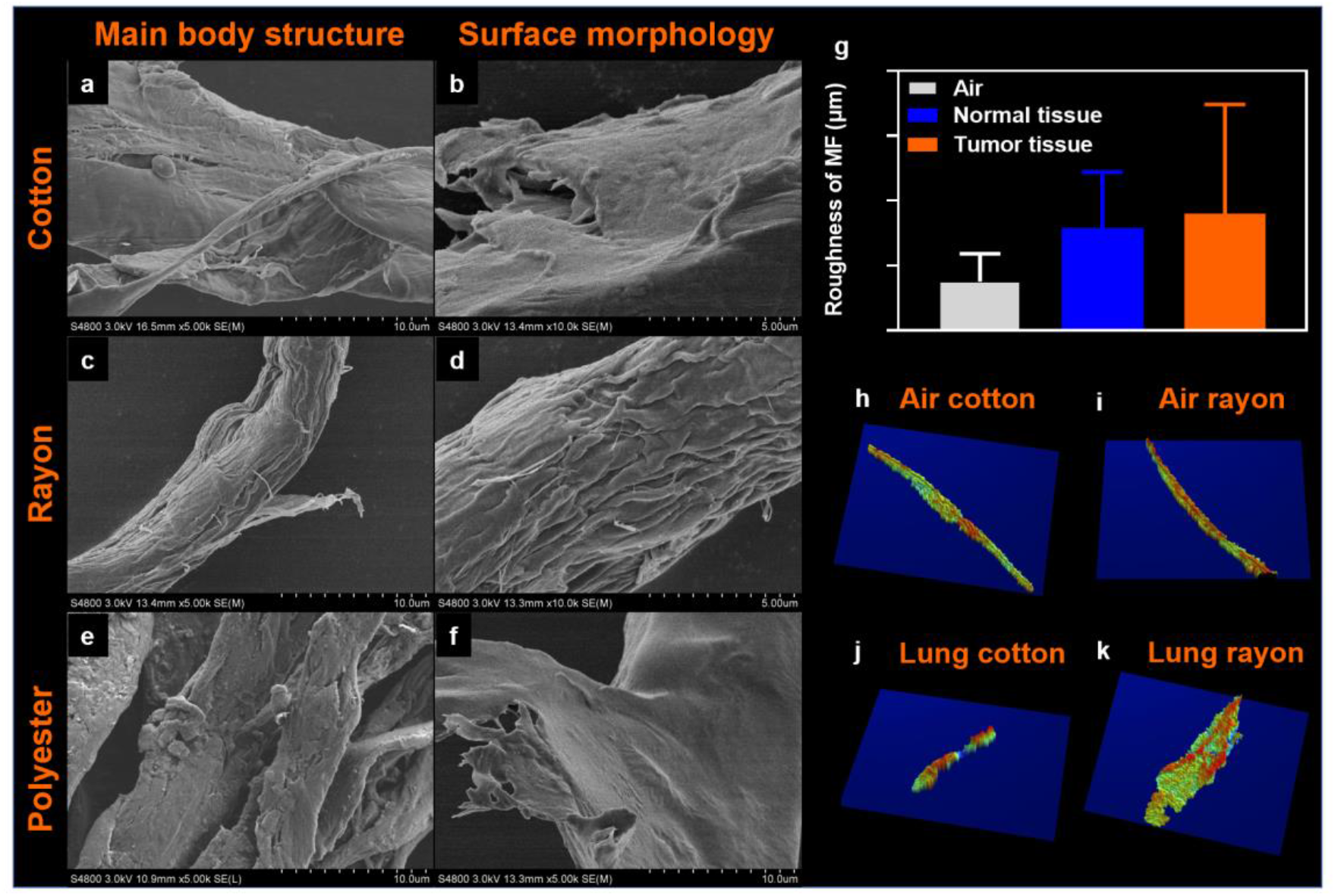
Representative pictures of microfibers isolated from lung tissues under scanned electron microscope (SEM). (a) main body structure of cotton; (b) surface morphology of cotton; (c) main body structure of rayon; (d) surface morphology of rayon; (e) main body structure of polyester; (f) surface morphology of polyester; (g) the roughness value (Ra) for microfibers from air deposition, normal tissues and tumor tissues (*n*=4 with 2 cotton microfibers, 1 rayon, and 1 polyester microplastics for each group according to their detected composition percentages); (h) representative surface roughness images for cotton microfiber from air deposition; (i) representative surface roughness images for polyester microplastic from air deposition; (j) representative surface roughness image for cotton microfiber from lung tissues; (k) representative surface roughness image for rayon microplastic from lung tissues.

## Discussion

The interrogation of the correlation of microplastics with diseases is an emerging field. The presence of microplastics in human tissues including gastrointestinal tract and placenta^34, 35^ has been previously reported. In this study, we provided solid evidence demonstrating the presence of microplastics in human lung tissues. Preliminary data demonstrated a higher detection rate of microplastics in the GGNs in comparing to the adjacent normal lung tissues. Moreover, the history of exposure to microfibers correlates with the occurrence of GGNs, which implicates an involvement of microfiber exposure in the etiology of GGNs.

### High abundance of microfibers and microplastics in human lung

The average wet weight of tissue samples used in this study is 44±32 mg, with 0.65±0.70 microfiber/sample detected. Thus, the predicted microfiber abundance in a whole human lung can reach 11,818 microfiber/lung (calculated as 800 g). As the content of microplastics accounting for 45% and 33% in normal and tumor tissues respectively, it is predicted that microplastic abundance in a whole lung can be 3,900-5,318 microplastic/lung. In our study, we found that the detection rate of microfibers in normal tissue is significantly increased with age; however, the detection rates of microfibers in patients with GGN of all ages are equivalent and high. This phenomenon indicates that microfibers may affect the occurrence of GGN after reaching the cumulative threshold.

The microfibers detected in the lungs were mainly ranged from 1000 to 5000 μm in length. This is different from the length distribution of microfibers in mountainous areas or medium-sized cities. For instance, a recent study found only around 11% of atmospheric deposited microfibers ranged in >1000 μm collected from a remote mountain catchment (French pyrenes)^36^. In the medium-sized Dongguan city, atmospheric fallout microfibers longer than 1200 μm constituted around 44%^37^. However, our microplastics distribution is close to the atmospheric microplastics distribution in an urban area of Paris, with 49% fell in the range of 1000-5000 μm^38^. This suggests that large cities may have a high proportion of long fibers in the air than medium-sized cities or remote areas. Of note, since our patients come from different cities and rural areas in China, the high percentage of long fibers in lungs may be because the longer inhaled microfibers may be easily entangled in lungs and difficult to be dispelled.

In addition to artificial fibrous microplastics, the cotton microfibers occupied a high proportion (39% in tumor tissue and 52% in normal tissue). Due to the not easily biodegradable nature, these cotton microfibers should have similar toxic effects as microplastics, if the indirect toxic effects of microplastics by releasing additives is not taken into account.

Most of the pulmonary microfibers probably come from indoor or outdoor air exposure, because the types of microfibers in lung tissue are highly similar to those found in the atmospheric fallout^11^, with the highest content being cotton, rayon and polyester. Previous studies have highlighted the presence of synthetic fibers in the lung tissue of workers in the textile industry, showing cases of respiratory irritation^39, 40^. The inhalability of a particle is size and shape dependent, as only the smallest particles below 5 µm and fibrous particles seem to be able to be deposited in the deep lung. Even though most of the bigger particles (inhalable particles) are subjected to mucociliary clearance in the upper airways, some of them can escape this mechanism and also be deposited in the deep lung. These particles (especially the longer fibers) tend to avoid clearance and show extreme durability in physiological fluids, likely persisting and accumulating when breathed in. This may be the reason why the proportion of long fibers (>1000 μm) in lung tissue (48-63%, **Figure S1**) is much higher than that in urban (44% for >1200 μm)^37^ or mountainous air (11% for >1000 μm)^36^. Previous studies report microfiber lengths merely. In this study, we firstly report the microfiber width (or diameter) dimension of microfibers in lung tissues, which is vital factor regulating the deposition of microfibers in the respiratory tract^41^.

The morphology of the inhaled fibers (e.g., air pollutants and microplastics) may affect their aerodynamic properties^42, 43^ which thereby finally modulate their ability to deposit in the distal pulmonary acinar airways as a result of varied width. Width is the main factor that determines whether or where in the respiratory tract that a microfiber can deposit^41^.Our results suggest that 13-125 μm wide microfibers/microplastics can reach lung tissues, with a mean width value of ∼30 μm. Once deposited, fiber length is the main factor that determines whether a microfiber can be effectively cleared^41^. Such elongated fibers (a mean value of ∼1.4 mm) tend to align with respiratory airflows, due to higher drag forces that resist sedimentation under gravity, and thus present a special clearance challenge for the lung compared with spheres of equivalent mass^44^. The thin, elongated shape of microfibers also affects clearance rates, enabling them to get entrained into the epithelial layer while inhibiting engulfment by macrophages ^41, 45^. Combined with a high surface-to-volume ratio compared to spheres of equivalent mass, such characteristics render non-spherical particles, and fibers in particular, promising candidates to be delivered to the deep lungs, which is supported by the finding that the microfibers identified in the lung are elongated in this study. Furthermore, the surface-to-volume ratio, hydrophobicity, size, and other properties of the microfibers can enhance their ability to attach heavy metal, increasing damage to the pulmonary immune microenvironment.

### Microfibers and the etiology of GGNs

Generally, when microfibers enter the respiratory system, the lung tissue has two main ways to dispel them. If microfibers deposited on the surface of respiratory mucosa, they can be excreted out of the respiratory tract by coughing or mucociliary escalator^46^. If microfibers reached the alveolar region, macrophages in the alveoli could remove the microfibers through phagocytosis, migration, or lymphatic transportation mechanisms^47^. Macrophages in alveoli often have high clearance efficiency for particles larger than 1 μm, thus microfibers at the micro-sized level are theoretically difficult to remain in lung tissue^48^. However, if the lung is continuously exposed to microfibers for a long time, it may cause excessive secretion of chemokines in the lung, thus destroying the normal chemotactic gradient, and may result in macrophages containing ingested microfibers resting in the alveolar space. Or if macrophages engage in “frustrated” phagocytosis for microfibers, digestive enzymes and other cellular contents will be spilled into the alveolar space, and this may mount innate immune responses which drive the sterile inflammation, fibrosis and other malignancies in the lung^43^, which thereby participate in the formation of GGNs and other inflammatory respiratory diseases.. The ingestion of microfibers by macrophage may mount innate immune responses which drive the sterile inflammation in the lung, which thereby participate in the formation of GGNs and other inflammatory respiratory diseases. Moreover, the interaction between vitreous particles/fibers and cells may lead to lung inflammation via intracellular messengers and cytotoxic factors which are released, and then cause secondary genotoxicity due to the continuous formation of reactive oxygen species^47, 49^.

Increasing evidence demonstrated that microfiber carry bacteria/virus^50^, which may play an important role in their interaction with the host. Intriguingly, compared to normal tissue, GGNs harbored a distinct lung bacterial community structure, with significantly increasing Firmicutes and Bacteroidetes ^51-53^. Even though there is no report about the biofilm community on airborne microplastics, but the biofilm community structures on microplastics are more affected by morphology/surface texture rather than polymer composition^54^. Therefore, we can further perform microbiota analysis on GGNs and normal tissues containing microfibers to further reveal the influence of microfiber on the lung microbiota. It’s tempting to speculate whether the microbes carried along by microfibers play a role in shaping the specific structure of microbiota in GGN. The microbiota analysis of GGNs and normal tissues that contain or free of microfibers will facilitate to understand the role of microfiber in modulating lung microbiota.

Moreover, airborne microplastic is an ideal vehicle for carrying micropollutants adsorbed to their hydrophobic surface, especially when related to urban environments. Recent studies performed using pre-diagnostic serum samples suggest that environmental exposure to micropollutants play an important role in the development of a series of cancers including lung cancer, prostate cancer, breast cancer, liver cancer and acute myeloid leukemia^55-58^. It’s therefore reasonable to speculate that microplastic may affect the occurrence of GGNs by delivering micropollutants which exerts toxic effect on the host. Of note, in addition to the adsorbed pollutants, microplastics may also contain unreacted monomers, additives, dyes, and pigments which could lead to adverse health effects^59, 60^. Though no heavy metal residue was found here by EDS (**Figure S4**), which may be due to the high detection limit of this method. However, there is no doubt that the high hydrophobicity and adsorption properties of microplastics will lead to the possibility of carrying pollutants into lung tissues, which needs further attention in the future.

### Surface roughness—a characteristic of microfibers awaits attention

Parameters commonly used to describe the characteristics of microfibers include polymer composition, size, color, density, shape, etc.; whereas little attention is paid to the index of surface roughness. Here we found all microfibers had obvious surface wear in both normal or tumor tissues. This kind of morphology is different from that found in the air, which shows both worn or unworn morphology and may be related to their aging degree in the environment. Besides, our pilot experiment confirmed that the increased surface roughness is not caused by the sampling, digestion and identification processes. It’s therefore highly likely that the surface roughness of microfibers was formed after entering the lungs. Some environmental processes, such as mechanical erosion by wind, water, or sand, UV radiation, biodegradation will increase the roughness of microplastics^5, 61, 62^. However, there is no report about the increase of roughness after microfibers/microplastics enter tissues. We believe this is the first report of the serious roughness phenomenon of microfibers in human tissues.

Little is known about the consequence of enhanced surface roughness to its interaction with the host. It would be interesting to check whether such characteristic of microfibers modulates its interaction with alveolar macrophages in multiple processes such as phagocytosis, recognition and mounting of immune responses. The types of cell death including apoptosis and necrosis in response to exogenous stimulation often show distinct effect on the outcome of diseases^63^. It would be fascinating to take the surface roughness as an important characteristic of microfibers into account in study its role in modulating the fate of macrophages and shaping of the microenvironment, which may constitute a new direction in future research of microfiber-host interaction. The technological advancement in acquisition of nano-scale microfibers will also promote the research on microfiber-host interaction.

### Microfibers link pulmonary GGN to occupational disease

Some occupational diseases may be related to long-term exposure to microfibers or microplastics. The “fiber-drawing workers” experienced a statistically significant excess in mortality from lung cancer^39^. Some of the nylon flock exposed workers had abnormal chest radiographs five-fold than non-exposed ones^40^. In the current study, we found that the microfibers significantly elevated in the normal lung tissues of pulmonary GGN patients with the increase of age. This phenomenon pinpoints the importance of the cumulative threshold of microplastics present in the lung in affecting the occurrence of GGN. Moreover, the data demonstrated that the long-term exposure to microfibers or microplastics is a risk factor for the formation of GGNs, indicating that it’s important for the clinicians to consider occupational exposure in the diagnosis of pulmonary GGNs. Our work also suggests that it’s important to perform the routine health examination in those people who have high risk to expose to microplastics, especially in the young people.

## Methods

### Clinical samples

Patients who underwent VATS lobectomy/sublobectomy for NSCLC in Shanghai Pulmonary Hospital between January 2020 and December 2020 were reviewed. Included in our studies were lung specimens with ground glass nodule (GGN). Tissue wet weight (0.043 ± 0.031 g, *n* =100). All patients received preoperative workups, including chest computed tomography (CT), brain magnetic resonance imaging (MRI), whole-body PET-CT and bronchoscopy. If N2 disease was suspected in PET-CT, endobronchial ultrasound was conducted to rule it out. This study was conducted in accordance with the principles of the Declaration of Helsinki. The study protocol was approved by the ethics committee of the Shanghai Pulmonary Hospital (IRB NO. K21-020), and informed consent from individual patients was waived for this retrospective analysis.

### Sample Digestion

Each lung tissue was transferred by stainless tweezer into a 40-mL glass tubes, and 30 mL of H2O2 (30 %, v/v) were added. The tubes were then incubated at 65°C and 80 rpm for 72 h for digestion. Afterwards, the digestate was filtered through 5 μm filter membrane (MCE, Millipore SMWP04700) and the filter membrane was stored in a dry glass petri dish for further observation and identification. All containers and tools were rinsed with Milli-Q water three times before use to avoid microfiber contamination. Procedural controls were conducted in parallel after the collection of lung tissues and throughout the following experiments.

### Observation and Identification of Microfibers

We used a Carl Zeiss Discovery V8 Stereo microscope (Micro Imaging GmbH, Germany) and an AxioCam digital camera to observe and photograph the substances on the surface of the membrane. Then, the composition of the suspected microfibers (including microplastics) was identified with μ-FT-IR (Nicolet iN 10, Thermo Fisher) under the transmission mode. A resolution of 4 cm^-1^ with a 16^-s^ scan time was chosen for data collection. All spectra were matched with our modified database and the result was accepted only when the matching index ≥ 70%. The Image J software (NIH Image) was used to measure the basic parameters (i.e. length, width and color) of fibers and fragments. Compare the colors of fibers and fragments with Pantone International Color Card by visual inspection, and express their colors with International standard color numbers and color scales.

### SEM/EDS analysis

To understand the surface morphology and elemental composition of microfibers or fragments from lung tissues, twelve representative samples (6 each of normal and tumor tissues) were studied using scanning electron microscope (SEM) (S4800) and Cryo-SEM (Lecia EM ICE, AFS2, FC7, TIC3X *, German) combined with energy-dispersive X-ray spectroscopy (EDS).

To confirm the effect of transportation, digestion, observation and identification processes on the surface morphology of microfibers, we conducted a pilot experiment by preparing two kinds of microfibers with the highest detection rate in our study (cotton and crayon) from the laboratory before and after all the above processes. Microfibers collected from indoor air in Shanghai were also collected for observation, including nine microfibers (3 each of cotton, rayon and polyester).

Microfibers were fixed on double-sided adhesive carbon tabs on the sample stage and spray gold. High resolution imaging was carried out by field emission SEM working at 3.0 KV and 15 μA, and the samples were taken at a magnification of 5.00 K. Qualitative elemental composition of six selected samples were determined by EDS working at 20.0kV and 20 μA.

### Microfiber surface roughness measurement

The surface roughness profile of microfibers was obtained by using a three-dimensional white light interferometery optical profiler Bruker Contour GT-K. The surface of middle area of microfibers were selected for detection. The entire profile data points were recorded, and the roughness average (Ra) of the absolute values of profile heights over a given area^64^ were calculated for each microfiber to evaluate their roughness, according to the standard method ASME B46.1-2009^65^.

### *In situ* Observation of Microfibers

The surgical lung tissues were cut into 30 μm slices and left untreated or treated with proteinase K (20 mg/mL) at 37 °C for 10 min. Each sample was then fixed to the microscope slide of dimension 25 mm × 75 mm using optical adhesive. To identify the *in situ* presence of microfibers in lung tissues, we used the Agilent 8700 Laser Direct Infrared (LDIR) Chemical Imaging System, equipped with a quantum cascade laser (QCL) source and a single point Mercury Cadmium Telluride (MCT) detector, at Agilent Technologies Application Laboratory in Shanghai, China. Briefly, the slides were firstly put in the LDIR system, and the height of samples were automatically identified and the tissue sections were focused. Then, the LDIR analyzer rapidly scanned the sample area at 1200 cm^-1^ with 20-μm resolution, and found and targeted areas containing suspect microfibers. Next, the targeted areas were enlarged and scanned with a fine resolution of 10 μm. Finally, the spectrum data were collected, and microfiber matching results were provided by conducting a library searching.

### Clinical indices

Through interviews with patients, we obtained first-hand information about the history of microfiber exposure of the patients. (1) We define people who smoke or have had a history of smoking as high risk of microfiber exposure, because cigarette butts contain various types of microfibers^66^ and could be inhaled in with the smoke. (2) We define people who have a high cooking frequency at home as high risk of microfiber exposure, because long time cooking and other housework will increase the microfiber exposure^67,68^. (3) We also inquired about the occupational background of the patients, and listed the patients who work in shoe factories, farms, construction sites, garbage cleaning, etc. as having high risks of microfiber exposure.

### Statistical Analysis

The microfiber or microplastic amount differences among tumor and normal tissue groups were evaluated by paired *t*-test. The microfiber exposure history and gender effects on the microfiber or microplastic detection results were evaluated by Chi-square test. All the above analyses were performed with SPSS (SPSS, version 20.0). Figures were plotted with GraphPad Prism (GraphPad, version 8.0).

## Data Availability

All data used during the study appear in the submitted article.

## Acknowledgements

We thank Professor Stefan HE Kaufmann (MPIIB, Berlin, Germany) for critical reading of the manuscript and insightful advice. We thank Jingjing Wang (Application Engineer of the Agilent Technologies, Shanghai, China) and Jinju Pei (Technical Engineer of the Agilent Technologies, Shanghai, China) for assistance with the *in situ* detection of microfibers in the lung slice. This project was supported by grants from the National Natural Science Foundation of China (81922030, 42077371 and 81770006 to H.L. and Q.C.). H. L. is sponsored by Projects supported by the National Science Foundation for Excellent Young Scholars of China (81922030), Shanghai ShuGuang Program (20SG19), Shanghai Pujiang Program (16PJ1408600) and Shanghai Medical and Health Services Outstanding Youth Talent Program (2017YQ078).

## Author Contributions

Q.C., C.C. and H.L. conceived the study and designed the experiments. Q.C., J. G., H.L. analyzed the data and wrote the original draft.

H. H. reviewed and edited the manuscript.

H.Y. performed the microfiber separation and characterization.

Y. Y. carried out the SEM/EDS observation.

Y.Y. and Q. C. participated in the roughness measurement.

Q. C. and J.G. carried out the LDIR measurement together with Agilent Co.

J. G., H. S., Y. C., and Y. R. collected the clinical samples.

## Ethics declarations

This study was conducted in accordance with the principles of the Declaration of Helsinki. The study protocol was approved by the ethics committee of the Shanghai Pulmonary Hospital (IRB NO. K21-020), and informed consent from individual patients was waived for this retrospective analysis.

## Data availability

The data are provided in the Figures and Supplementary Figures. Related information will be provided upon reasonable request.

## Figure Legends

**Figure S1.**
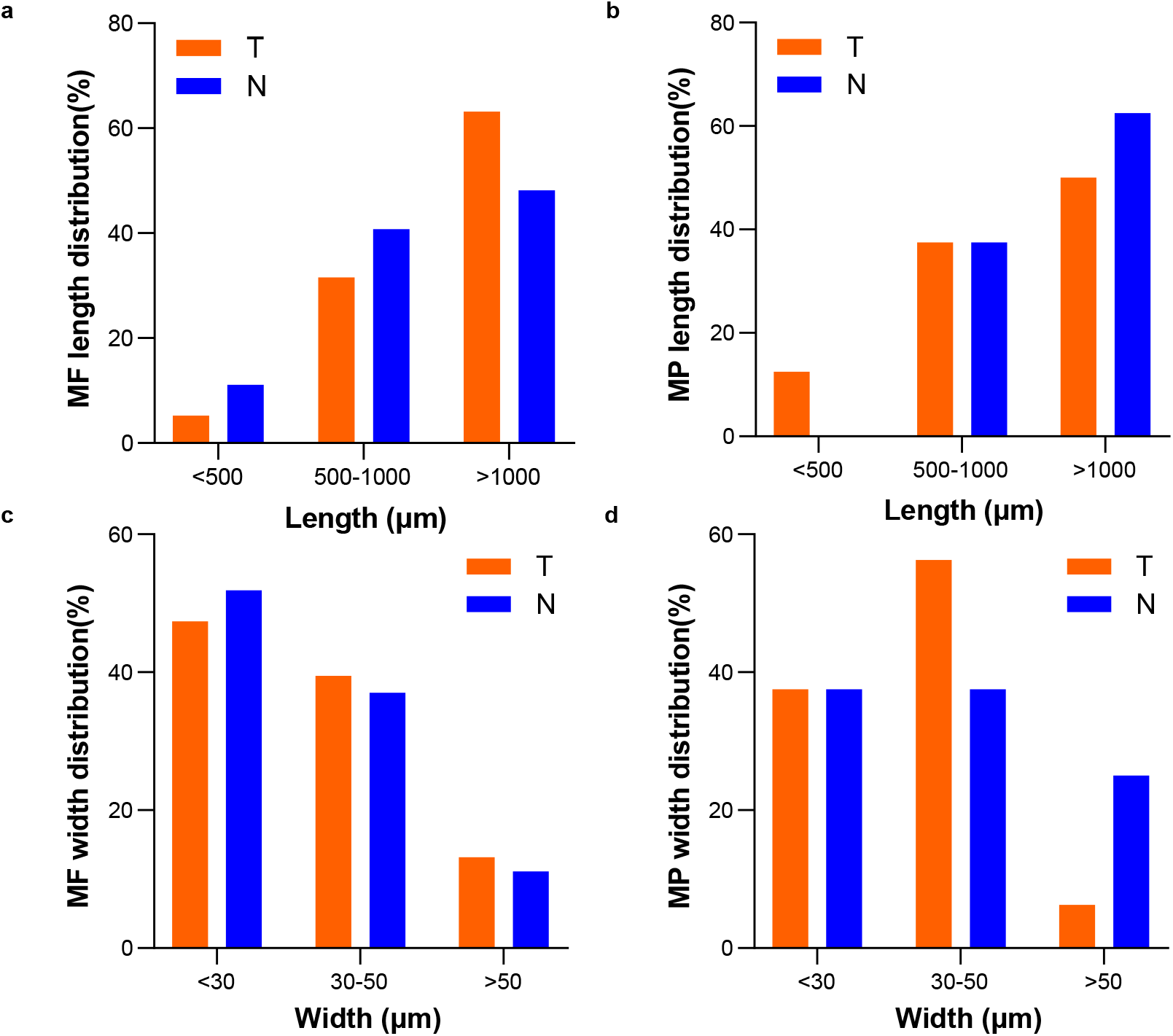
Length and width distribution of microfibers and microplastics. (a) the length distribution of microfibers; (b) the width distribution of microfibers; (c) the length distribution of microplastics; (d) the width distribution of microplastics. MF: microfiber; MP: microplastic.

**Figure S2.**
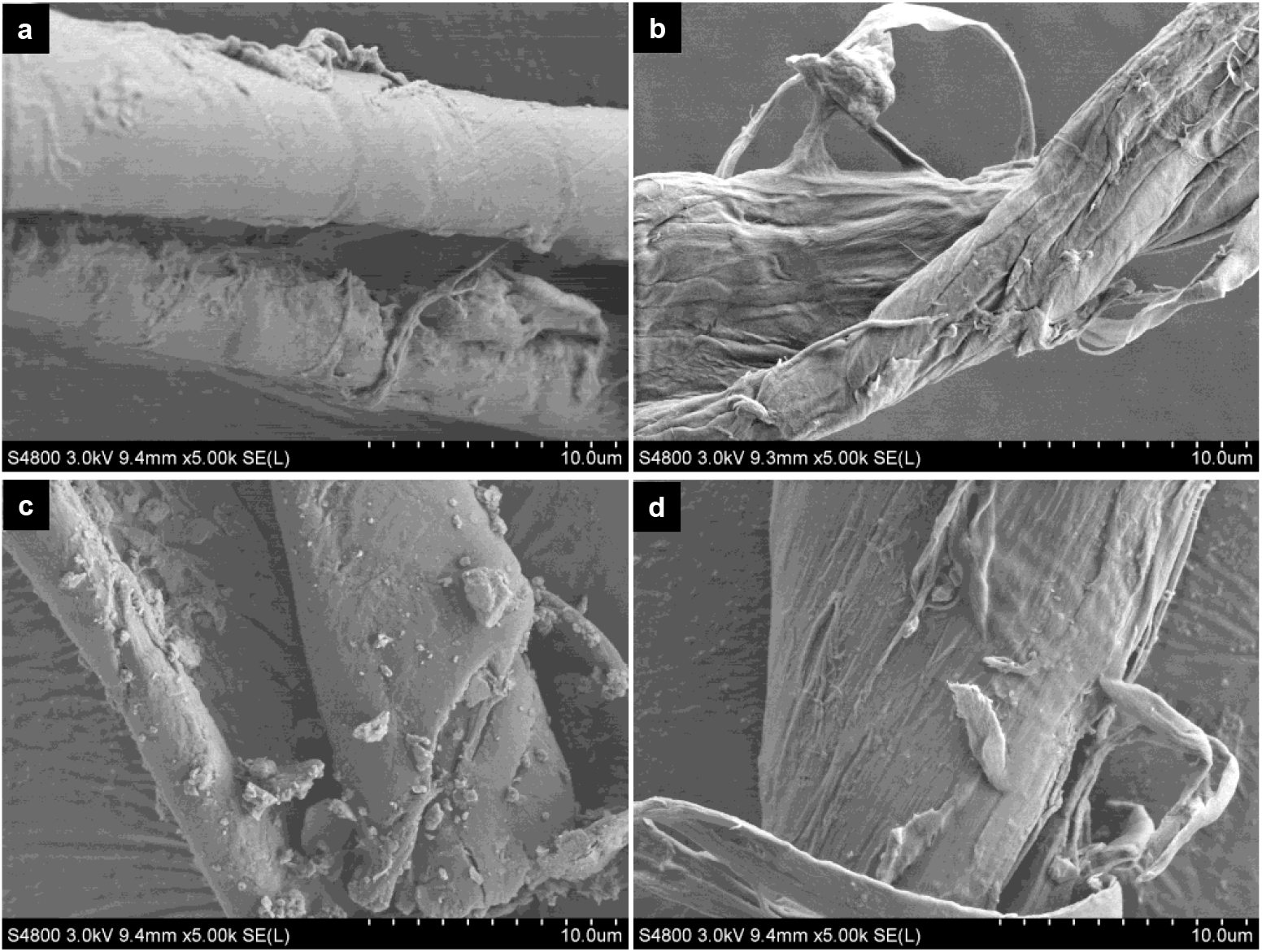
Representative pictures of airborne cotton and rayon. (a) cotton with smooth surface; (b) cotton with rough surface; (c) rayon with smooth surface; (d) rayon with rough surface.

**Figure S3.**
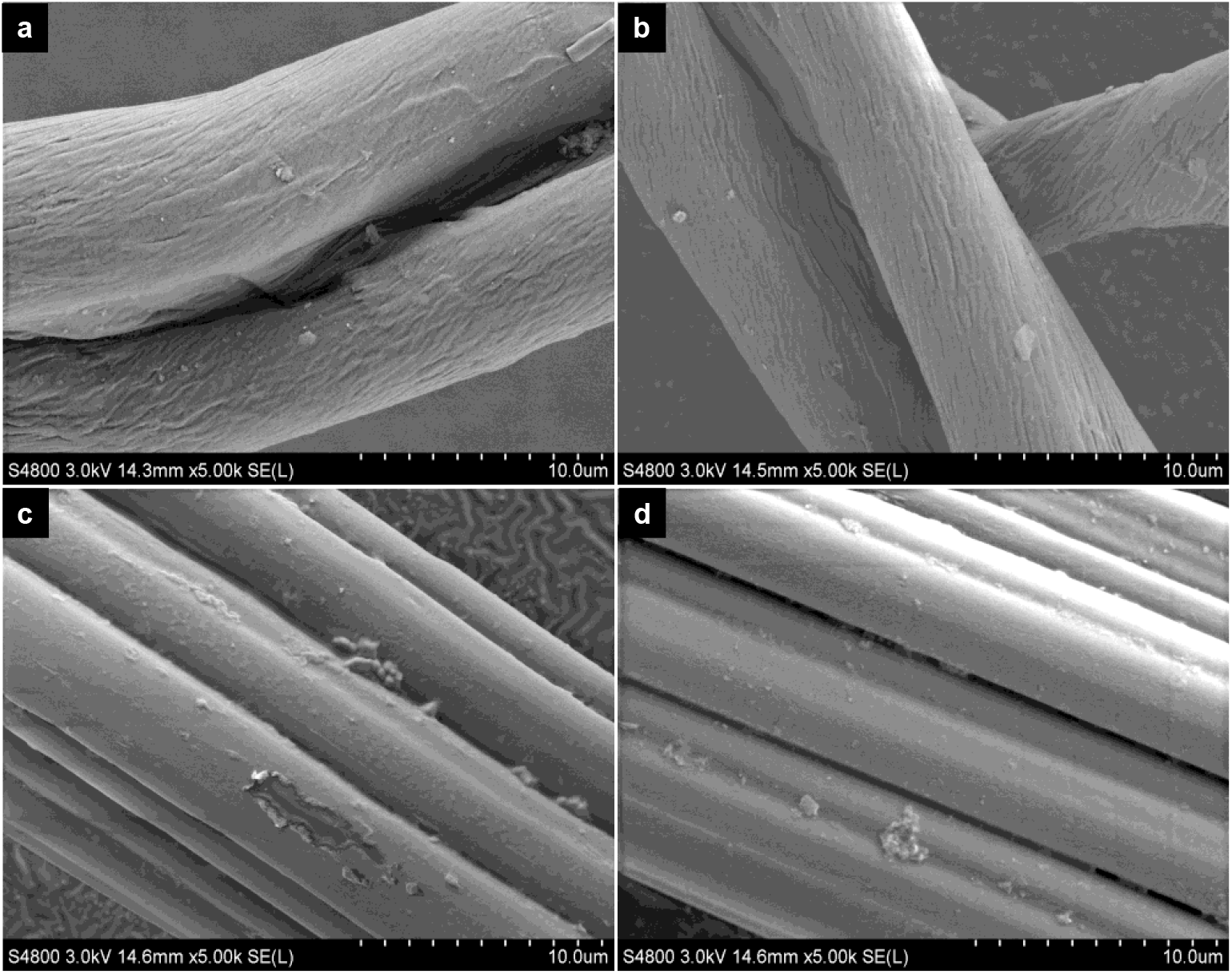
Representative pictures of laboratory prepared cotton and rayon microfibers before and after experimental processes (digestion, observation, and identification). (a) cotton before processes; (b) cotton after processes; (c) rayon before processes; (d) rayon after processes.

**Figure S4.**
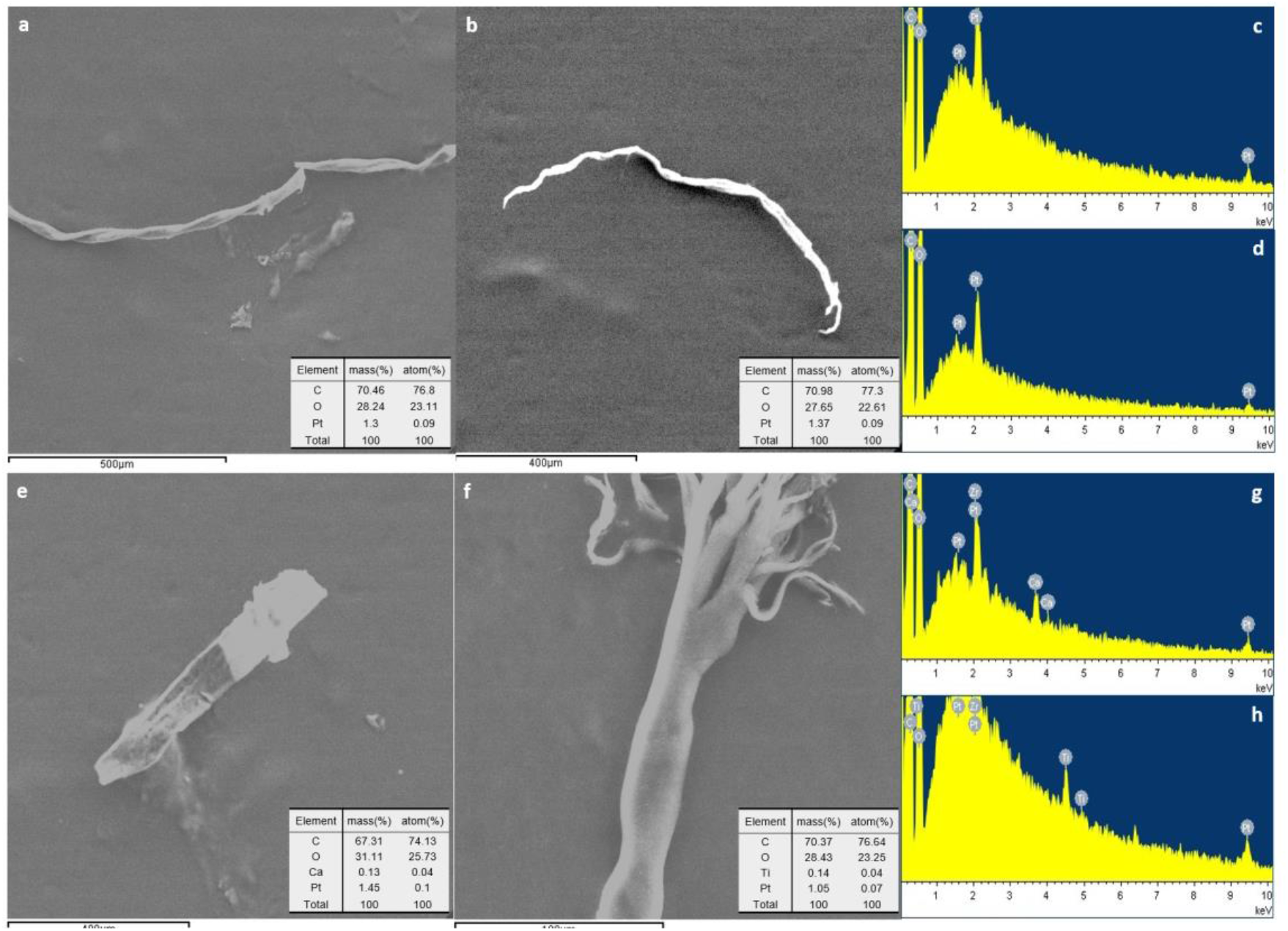
Microfiber surficial elemental composition illustrated by SEM/EDS. (a) cotton from a tumor tissue; (b) cotton from a normal tissue; (c) EDS figure for (a); (d) EDS figure for (b); (e) polyester from a tumor tissue; (f) polyester from a normal tissue; (g) EDS figure for (e); (h) EDS figure for (f).

**Table 1.** Clinical information of patients.

| No. | Sex | Age range | Tobacco exposure | Cooking | Microfiber exposure | Risk assessment |
| --- | --- | --- | --- | --- | --- | --- |
| 1 | Female | 45-64 | No | No | No | L |
| 2 | Male | 45-64 | No | No | No | L |
| 3 | Female | 45-64 | No | Yes | No | H |
| 4 | Female | NA | No | No | No | / |
| 5 | Female | 45-64 | No | No | Yes | H |
| 6 | Female | NA | No | No | No | / |
| 7 | Male | 45-64 | Yes | No | No | H |
| 8 | Female | 45-64 | No | No | No | L |
| 9 | Female | 45-64 | Yes | No | No | L |
| 10 | Male | 45-64 | Yes | No | No | H |
| 11 | Female | 65+ | No | No | No | L |
| 12 | Male | 65+ | Yes | No | Yes | H |
| 13 | Female | 25-44 | No | No | No | / |
| 14 | Male | 45-64 | Yes | No | Yes | H |
| 15 | Female | 45-64 | No | Yes | No | H |
| 16 | Female | 65+ | Yes | Yes | No | H |
| 17 | Female | 45-64 | No | Yes | No | H |
| 18 | Female | 65+ | No | No | No | / |
| 19 | Female | 45-64 | No | Yes | No | L |
| 20 | Female | 65+ | No | Yes | No | H |
| 21 | Female | 45-64 | No | Yes | No | H |
| 22 | Female | 45-64 | Yes | Yes | Yes | H |
| 23 | Male | 45-64 | Yes | No | No | H |
| 24 | Female | 45-64 | No | Yes | No | H |
| 25 | Female | 45-64 | No | No | No | / |
| 26 | Female | 45-64 | No | Yes | No | L |
| 27 | Male | 45-64 | No | No | No | L |
| 28 | Male | 65+ | Yes | No | Yes | H |
| 29 | Female | 45-64 | Yes | Yes | No | H |
| 30 | Female | 45-64 | Yes | Yes | Yes | H |
| 31 | Male | 45-64 | Yes | No | No | H |
| 32 | Female | 45-64 | Yes | Yes | No | H |
| 33 | Female | 45-64 | No | No | No | L |
| 34 | Male | 45-64 | No | No | No | L |
| 35 | Male | 65+ | Yes | No | Yes | H |
| 36 | Female | 25-44 | No | No | No | L |
| 37 | Female | 45-64 | No | No | No | L |
| 38 | Female | 45-64 | Yes | Yes | No | H |
| 39 | Male | 45-64 | Yes | No | No | H |
| 40 | Male | 45-64 | No | No | No | L |
| 41 | Male | 25-44 | No | No | No | L |
| 42 | Female | 45-64 | No | No | No | L |
| 43 | Male | 45-64 | Yes | No | Yes | H |
| 44 | Female | 25-44 | Yes | Yes | No | L |
| 45 | Female | 45-64 | No | Yes | No | H |
| 46 | Female | 25-44 | Yes | Yes | No | L |
| 47 | Female | 25-44 | No | No | No | L |
| 48 | Female | 45-64 | No | No | No | L |
| 49 | Female | 45-64 | No | Yes | Yes | H |
| 50 | Male | 45-64 | No | No | No | L |

**Table 2.** Characteristics of patients.

| **Characteristic** | **Patients(n=50)** |
| --- | --- |
| Female sex (n, %) | 34 (68.0) |
| Age (mean ± SD) (y) | 55.4±10.1 |
| History of diabetes (n, %) | 4 (8.0) |
| Hypertension (n, %) | 13 (26.0) |
| COPD (n, %) | 0 (0) |
| Coronary artery disease (n, %) | 2 (4.0) |
| Stroke (n, %) | 1 (2.0) |
| Pulmonary embolism (n, %) | 0 (0) |
| History of tobacco exposure (n, %) | 19 (38.0) |
| Adjuvant chemotherapy (n, %) | 1 (2.0) |
| Adjuvant radiotherapy (n, %) | 0 (0) |
| Neoadjuvant therapy (n, %) | 0 (0) |
| Adjuvant Target therapy (n, %) | 0 (0) |
| Surgical laterality (n, %) |  |
| Left | 18 (36.0) |
| Right | 32 (64.0) |
| Intrapericardial Procedure |  |
| Yes | 0 (0) |
| No | 50 (100) |

*COPD: chronic obstructive pulmonary disease.*

